# A foundation model for mapping the phenomic and genetic landscape of cerebral small vessel disease biomarkers

**DOI:** 10.64898/2025.12.19.25342615

**Authors:** Weikang Gong, Fang Lan, Peng Ren, Hao Hu, Wenjing Su, He-Ying Hu, Qiong-Yao Li, Xiao-He Hou, Liang-Yu Huang, Quan Hao, Yong-Li Zhao, Yan Fu, Dan-Dan Zhang, Wei Zhang, Ze-yu Li, Mingchen Ma, Ruijia Xu, Tiansiyu Wen, Lu-yang Zhang, Shu-Yi Huang, Biying Lin, Lingshuang Han, Mei Cui, Jin-Tai Yu, Wei Cheng

**Affiliations:** Institute of Science and Technology for Brain-Inspired Intelligence and School of Data Science, State Key Laboratory of Medical Neurobiology and MOE Frontiers Center for Brain Science, Fudan University, Shanghai, China; Department of Neurology, Huashan Hospital, Fudan University, Shanghai, China; Department of Neurology, Qingdao Hospital, University of Health and Rehabilitation Sciences (Qingdao Municipal Hospital), Qingdao, China; Department of Neurology, Qingdao Municipal Hospital, Qingdao University, Qingdao, China; Department of Geriatric, the Affiliated Hospital of Qingdao University, Qingdao University, Qingdao, China; Department of Neurology and National Center for Neurological Disorders, Huashan Hospital, State Key Laboratory of Brain Function and Disorders, MOE Frontiers Center for Brain Science, Shanghai Academy of Natural Sciences (SANS), Shanghai Medical College, Fudan University, Shanghai, China; Fudan ISTBI—ZJNU Algorithm Centre for Brain-Inspired Intelligence, Zhejiang Normal University, Jinhua, China; School of Medicine, Zhejiang University, Hangzhou, China; Department of Radiology, Hangzhou First People’s Hospital, Hangzhou, China; Centre for Functional MRI of the Brain (FMRIB), Nuffield Department of Clinical Neurosciences, Wellcome Centre for Integrative Neuroimaging, University of Oxford, UK

**Author notes:** Correspondence to: Dr. Weikang Gong, School of Data Science, Fudan University, Shanghai, China, Prof. Mei Cui, Huashan Hospital, Fudan University, Shanghai, China, Prof. Jintai Yu, Huashan Hospital, Fudan University, Shanghai, China, Prof. Wei Cheng, Institute of Science and Technology for Brain-Inspired Intelligence, Fudan University, Shanghai, 200433, China. These authors contributed equally to this work (Weikang Gong, Fang Lan, Peng Ren, Hao Hu, Wenjing Su).

## Abstract

Cerebral small vessel disease (CSVD) is a leading cause of age-related cognitive decline and neurological disorders, yet its precise characterization in large populations has been constrained by reliance on subjective neuroimaging ratings. To address this, we developed CSVDtransformer, a foundation model that simultaneously quantifies six key CSVD biomarkers from structural brain MRI. In 3,718 subjects, the model achieved excellent accuracy (mean AUC = 0.904) in measuring periventricular and deep white matter hyperintensities, Fazekas scores, enlarged perivascular spaces, lacunar infarcts, and cerebral microbleeds. Validation across two independent, external datasets (N=568) confirmed its robust generalizability. As a clinical decision-support tool, it augmented neurologist assessment relative accuracy by 20%. Application to 59,772 UK Biobank participants revealed distinct associations of these quantified biomarkers with incident stroke, dementia, and psychiatric disorders. Large-scale multi-omics analysis identified 1,365 significant plasma protein correlates and 14 novel genetic loci for these CSVD biomarkers. These associations implicate pathways of endothelial dysfunction, inflammation, and lipid metabolism. Mendelian randomization analyses provided evidence for causal relationships between specific vascular-metabolic proteins and CSVD biomarkers, such as positive effect of EFEMP1 and negative effect of EPO on CSVD. Furthermore, drug-target enrichment analysis highlighted the potential for targeting TFPI and EPO to address vascular dysfunction associated with CSVD. Our study establishes CSVDtransformer as a scalable foundation model that deciphers the complex systemic biology of cerebral microvascular health.

## Introduction

Cerebral small vessel disease (CSVD) is among the most prevalent age-related brain pathologies and a major contributor to stroke, dementia, and cognitive impairments worldwide. Radiologically, CSVD is characterized by key MRI features including white matter hyperintensities^1^, enlarged perivascular spaces in basal ganglia^2^, lacunes, and cerebral microbleeds (CMB), that collectively reflect microvascular injury and disrupted brain integrity. These markers are highly prevalent in mid- to late-life and account for a substantial proportion of stroke and dementia burden globally^3,4^.

Beyond these traditional vascular outcomes, recent studies have increasingly highlighted that CSVD extends its impact beyond traditional vascular outcomes to include a broad range of cognitive impairments and neuropsychiatric disorders. For instance, CMB has found to be associated with worse cognitive function^5^. Higher WMH volumes and are associated with late-life depression, suggesting that CSVD may contribute to the pathophysiology of mood disorders^6,7^. In Alzheimer’s disease, WMH were found to predict amyloid increases in the early stages^8^. Similarly, in Parkinson’s disease, comorbidity with CSVD has been shown to exacerbate motor symptoms and cognitive impairment, highlighting the broader role of cerebrovascular health in neurodegenerative disease progression^9^. These findings collectively support the role of CSVD as a critical factor in the pathophysiology of neurodegenerative disorders.

Despite their clinical relevance, current quantification of CSVD biomarkers largely depends on visual rating or semi-manual segmentation, which is time-consuming, subjective, and poorly scalable for quantitating large-scale cohorts. Recent advances in artificial intelligence (AI) have begun to transform the field of neurovascular imaging. Machine-learning models trained on MRI can extract subtle, spatially distributed signatures of microvascular pathology, offering reproducible and high-throughput quantification of CSVD^10–14^. For instance, studies adapting models such as convolutional neural network (CNN) and transformer have demonstrated their ability to reliably quantify WMH in heterogeneous patient cohorts, outperforming traditional segmentation methods and showcasing their potential for large-scale clinical applications^10,12^. Similarly, studies using deep-learning models based on susceptibility-weighted imaging (SWI) have demonstrated better performance in detecting CMB than traditional methods and even neurologists’ assessments^13,14^. However, most existing AI studies have focused on single CSVD markers and are limited by small, annotated datasets (typically N < 1000), which constrains cross-scanner generalizability and the robustness of subsequent analyses. A comprehensive, accurate, and robust AI framework for the automated assessment of the full spectrum of CSVD biomarkers is currently lacking. Furthermore, the relationships between these automated biomarkers and cognitive, clinical, genetics and multi-omic profiles in large populations remain largely unexplored.

To address these gaps, we leveraged large-scale multimodal data from the UK Biobank (UKB), with expert annotations for approximately 4,000 subjects, the largest such dataset to date, to our knowledge. We developed CSVDtransformer, a novel architecture that utilizes a feature reranking mechanism to predict CSVD biomarkers from three structural MRI modalities (T1w, T2-FLAIR, and SWI). This approach moves beyond the spatial-locality bias of conventional models by leveraging the statistical correlation between each voxel and the CSVD biomarkers to form functionally coherent patches. This ensures the model prioritizes the most discriminative, and often spatially dispersed, imaging biomarkers, enhancing both data-efficiency and its ability to capture the holistic biomarkers of CSVD. We rigorously validated our model on internal and external datasets, comparing its performance against other state-of-the-arts and expert annotations. We also evaluated its utility as an assistive tool for improving diagnostic accuracy among junior clinicians. Finally, we automatically extracted CSVD markers across 59,772 UKB subjects to systematically investigate their associations with a wide range of cognitive, clinical, genetic, and metabolic markers. Our analyses aimed to (i) conduct phenome-wide association analyses linking CSVD biomarkers to diverse brain and systemic disorders; (ii) to identify genetic and metabolic signatures underlying CSVD biomarkers; and (iii) to provide a systems-level understanding of cerebrovascular health and disease vulnerability. Together, this framework integrates neuroimaging, multi-omics, and population neuroscience to delineate the biological landscape of cerebral microvascular brain health (Fig. 1).

**Fig. 1.**
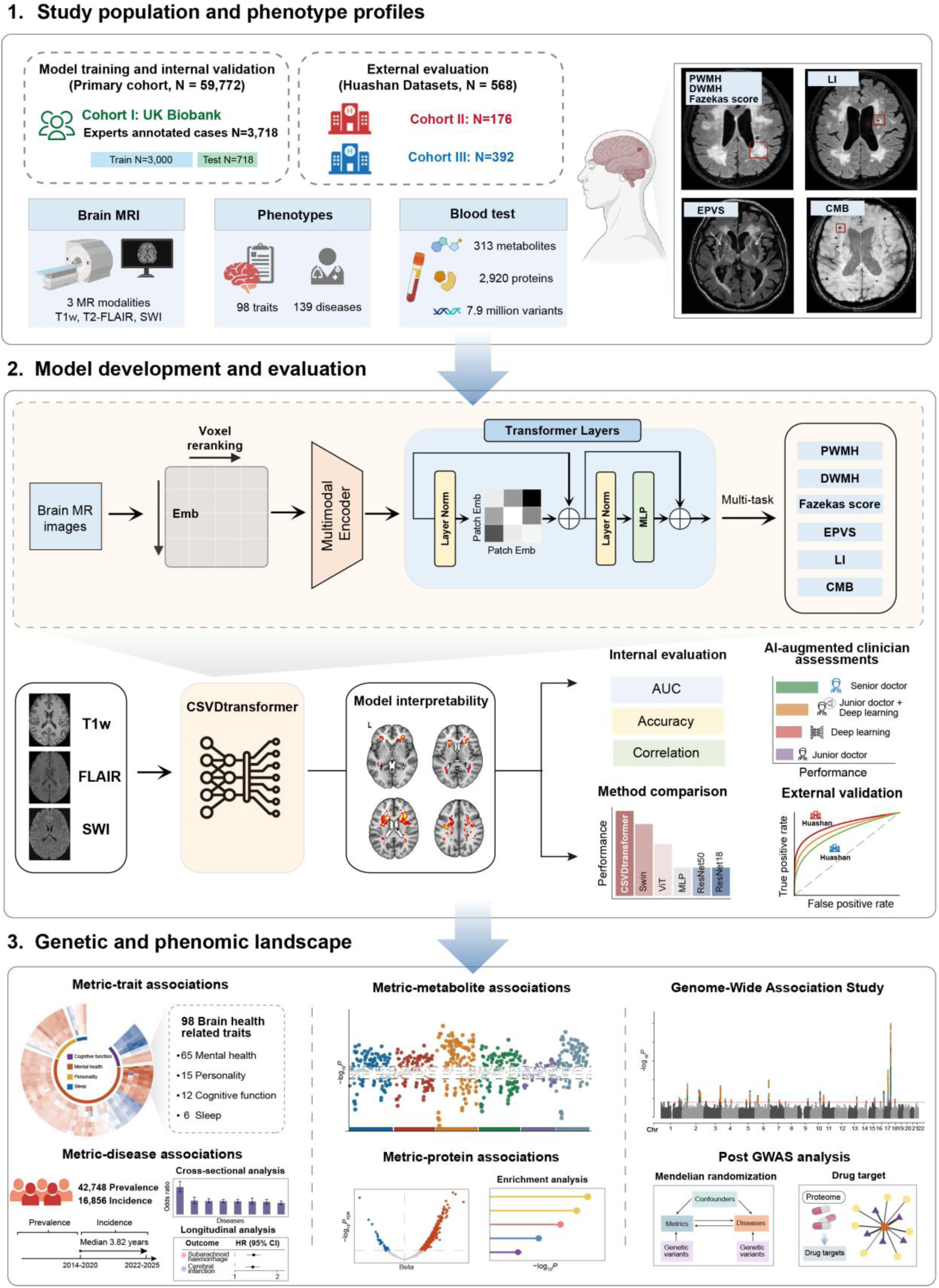
Study Overview. MR images illustration of CSVD biomarkers (WMH, LI, and CMB) were obtained from Ke et al.^89^. EPVS image was obtained from Potter et al.^90^.

## Results

### Cohort characteristics and CSVD biomarker quantification

Our study included 59,772 participants from the UKB imaging cohort after applying quality control exclusions for incomplete data (mean age 65.2 years; 55.8% female). The cohort was followed for a median of 3.82 years (interquartile range 2.72–5.20), with 138 incident neurological or psychiatric events recorded during this period.

To establish ground truth for model training and validation, a subset of 3,718 UKB participants was rigorously annotated by a team of eight physicians, each with 6 to 10 years of clinical experience. All annotations adhered to the standardized definitions for CSVD imaging markers set forth by the Standards for Reporting Vascular Changes on Neuroimaging (STRIVE). Subsequently, the CSVDtransformer model was trained and validated on this annotated cohort and then applied to the entire cohort to generate precise, quantitative measures for all six CSVD biomarkers.

### CSVDtransformer accurately quantifies diverse CSVD biomarkers

We developed CSVDtransformer, a foundation model to simultaneously quantify six key CSVD biomarkers from structural brain MRI: periventricular white matter hyperintensities (PWMH), deep white matter hyperintensities (DWMH), Fazekas score, enlarged perivascular spaces (EPVS), lacunar infarcts (LI), and cerebral microbleeds (CMB).

CSVDtransformer achieved excellent predictive performance across all biomarkers. For white matter hyperintensity-related features (PWMH, DWMH, and Fazekas score), the model attained a mean area under the curve (AUC) of 0.932. It also demonstrated high accuracy for other lesion types, with AUCs of 0.843 for EPVS, 0.857 for LI, and 0.934 for CMB (Fig. 2a).

**Fig. 2.**
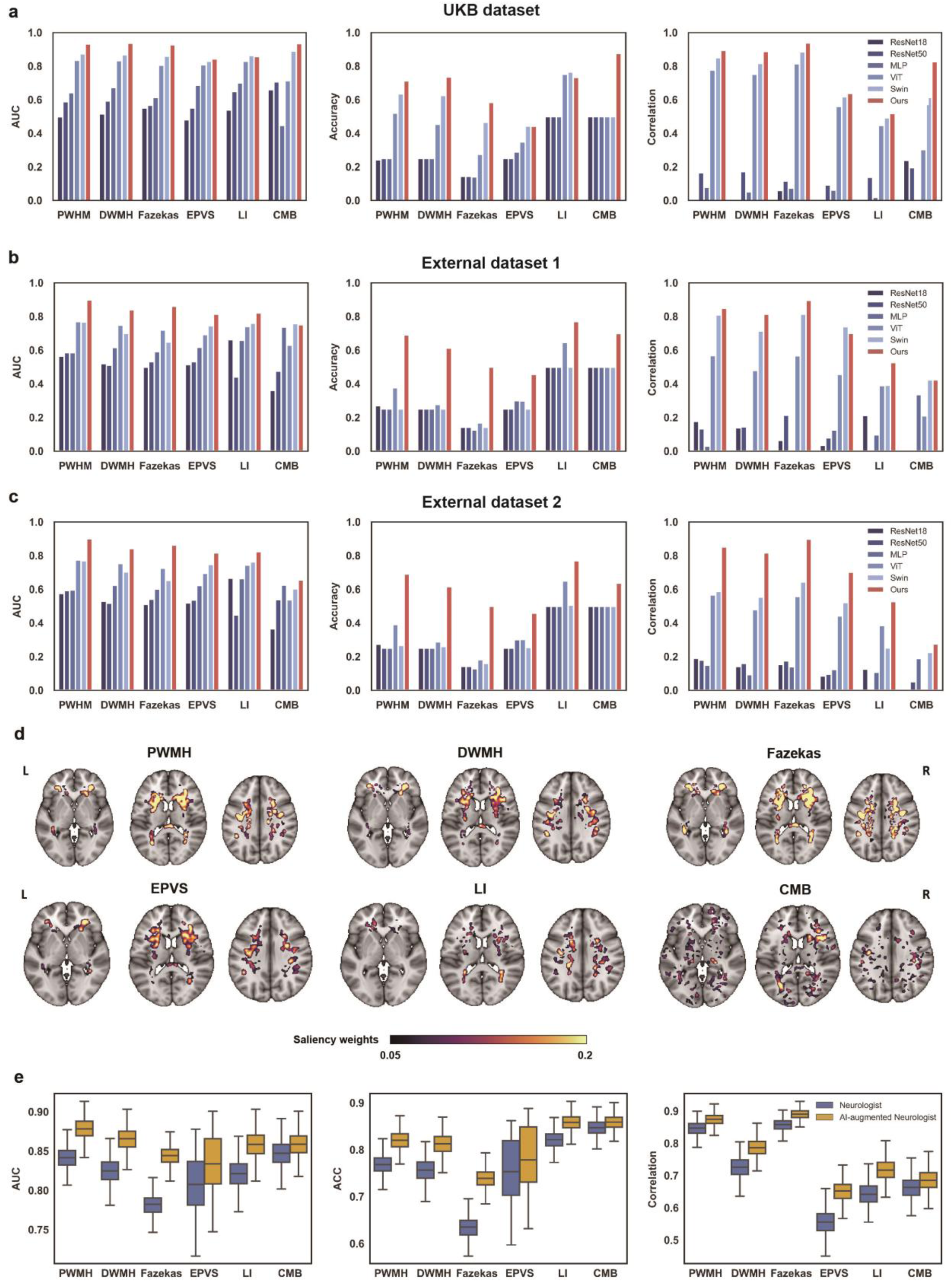
CSVDtransformer model performance and interpretability. (a) Model performance in the UK Biobank dataset. Bar plots show the area under the curve (AUC), accuracy, and correlation across six CSVD biomarkers (PWMH, DWMH, Fazekas score, EPVS, LI, and CMB). Performance of the proposed transformer-based model (red) is compared against benchmark models, including ResNet-18, Swin-Transformer, U-Net, and ConvNeXt. (b, c) External validation in two external datasets demonstrating consistent performance patterns across metrics and CSVD biomarkers. (d) Model interpretability maps derived from attention-weighted activation regions for each CSVD biomarker. The highlighted voxels (in red–yellow) indicate brain regions contributing most to model prediction, overlaid on the MNI152 template (L, left; R, right) (e) External validation of CSVDtransformer used as a decision-support tool improves the diagnostic accuracy of neurologist ratings for CSVD biomarkers.

In benchmark comparisons, CSVDtransformer substantially outperformed other state-of-the-art architectures. It exceeded the second-best model, Swin Transformer, a benchmark in brain imaging analysis^15^, by 24.6% in mean AUC. This performance advantage was consistent across multiple metrics, with an average improvement of 11.5% in classification accuracy and 14.1% in Pearson correlation over Swin Transformer. CSVDtransformer achieved greater margins over standard Vision Transformers and CNN-based models (Fig. 2a).

To evaluate the model’s robustness, we tested its generalizability on two external, independently annotated clinical datasets with patients having overall higher CSVD burden (N=568). Without any retraining, CSVDtransformer maintained robust performance despite shifts in scanner protocols and population characteristics. On External Dataset 1, the model achieved a strong mean AUC of 0.866 for WMH-related biomarkers, with individual AUCs of 0.814 for EPVS, 0.821 for LI, and 0.751 for CMB (Fig. 2b). On External Dataset 2, it attained a mean WMH AUC of 0.868, with AUCs of 0.816 for EPVS, 0.823 for LI, and 0.654 for CMB (Fig. 2c). Notably, in these challenging external settings, CSVDtransformer surpassed the second-best method by an average of 37.4% in AUC, underscoring its superior generalizability (Fig. 2b, c).

To validate that the model’s decisions were anatomically grounded, we employed interpretability methods to generate spatial saliency maps^16,17^. These maps confirmed that CSVDtransformer leveraged clinically meaningful neuroanatomy for its assessments (Fig. 2d). The model distinctly localized PWMH to periventricular regions and DWMH to deep white matter, while the Fazekas score integrated features from both. EPVS and lacunes were associated with white matter tracts connected to the basal ganglia, and CMB were correctly identified across diverse cortical and subcortical regions. This anatomical coherence confirms that the model captures fundamental radiological signatures of CSVD, although its attention may not always encompass all regions considered during expert manual annotation.

### CSVDtransformer enhances neurologist CSVD biomarker rating accuracy

We compared assessments made by practicing clinicians with those made by clinicians assisted by CSVDtransformer, using the Huashan dataset 2 (N=392) for six CSVD biomarkers. Model assistance consistently improved performance across all biomarkers, with notable increases in AUC, accuracy, and correlation (Fig. 2e). Relative to unaided clinician ratings, the mean AUC improved by 19.9%, accuracy by 19.9%, and correlation by 18.5% across the six biomarkers.

### Associations between CSVD neuroimaging biomarkers and health-related traits

Associations between the six CSVD biomarkers and a comprehensive set of brain health–related traits were examined (Fig. 3a; Supplementary Table 1). After false discovery rate correction (FDR < 0.05), a total of 325 significant associations were identified, comprising 62 significant associations from DWMH, 57 from PWMH, 56 from Fazekas score, 60 from CMB, 34 from LI, and 56 from EPVS.

**Fig. 3.**
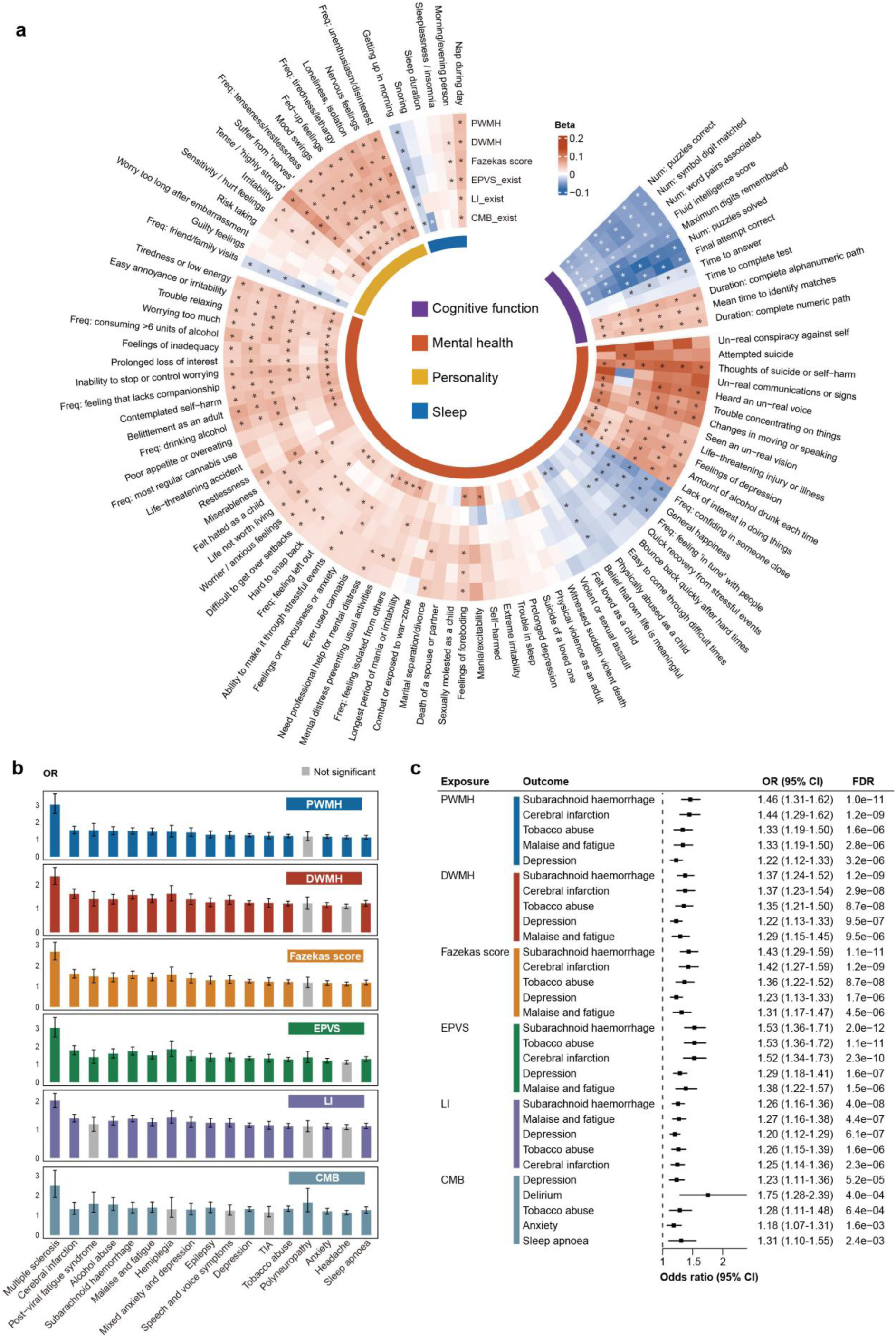
PheWAS analysis. (a) Associations between six CSVD biomarkers and 98 brain health–related traits. Abbreviations of each trait were described in Supplementary Table 2. (b) Cross-sectional disease associations between CSVD biomarkers and 16 neurological and neuropsychiatric disorders, restricted to diseases with ≥50 prevalent cases (*P*_FDR_ < 0.05). Bars represent odds ratios from logistic regression models adjusted for age, sex, education, and intracranial volume. (c) Longitudinal risk associations based on Cox proportional hazards models, showing the top five prospective disease outcomes for each CSVD biomarker. Error bars denote 95% confidence intervals. TIA, transient ischemic attack.

In the mental health and well-being domain, higher scores were related to depression, experience of adverse life events, anxiety and panic, self-harm behaviours, social situation, mental distress, psychotic experiences, alcohol consumption, and psychosocial factors such as feeling of miserableness. Among sleep-related traits, higher inference scores were associated with more frequent napping, increased snoring, less easy getting up in the morning, and evening sleep chronotype. Within personality measures, significant correlations were observed between CSVD biomarkers and increased frequency of tiredness/lethargy and unenthusiasm/disinterest, tense, mood swing, fed-up feelings, loneliness and isolation, nervous feelings, irritability, tiredness, as well as lower frequency of friend/family visit. Finally, in the cognitive domain, higher scores were associated with slower reaction time, lower fluid intelligence score, and poorer performance in symbol digit substitution, trail making, symbol digit substitution, paired associate learning, numeric memory, prospective memory, and matrix pattern tasks (all *P*_FDR_ < 0.05; Supplementary Table 2).

### Associations of CSVD biomarkers with neurological and neuropsychiatric disorders

To ensure stable model estimation, we restricted analyses to diseases with at least 50 prevalent cases in the cross-sectional analysis and at least 50 incident cases during follow-up in the longitudinal analysis. This resulted in 22 prevalent diseases and 36 incident diseases for inclusion. After FDR correction, cross-sectional analyses identified significant associations between CSVD biomarkers and 21 diseases, whereas longitudinal analyses identified significant associations with 18 diseases. (Fig. 3b, 3c; Supplementary Table 4 and 5).

In the cross-sectional analyses, the most prominent associations were observed for multiple sclerosis, which showed consistent effects across all six CSVD biomarkers (odd ratio (OR) range = 2.02 – 3.02, all *P*_FDR_ < 1 × 10^-26^). Other top associations included acute intoxication due to use of alcohol with EPVS (OR = 1.87, *P*_FDR_ = 1.32 × 10^-7^), hemiplegia with EPVS (OR = 1.83, *P*_FDR_ = 2.19 × 10⁻⁶), cerebral infarction EPVS (OR = 1.76, *P*_FDR_ = 4.36 × 10⁻¹³), and subarachnoid haemorrhage EPVS (OR = 1.72, *P*_FDR_ = 2.97 × 10^-15^). Additional associations were identified for polyneuropathy, alcohol abuse, post-viral fatigue syndrome, malaise and fatigue, epilepsy, speech and voice symptoms, anxiety, depression, sleep apnoea, and headache (all *P*_FDR_ < 0.05, Supplementary Table 4).

In the longitudinal analyses, hazard ratios (HRs) ranged from 1.13 to 1.75 across significant associations, based on a mean follow-up period of 3.77 years among a total of 49,817 participants. The most prominent associations were observed for delirium with CMB (HR = 1.75, *P*_FDR_ = 4.02 × 10^-4^). Other top associations included dementia (EPVS; HR = 1.69, *P*_FDR_ = 1.06 × 10^-5^), emotional state symptoms (EPVS; HR = 1.62, *P*_FDR_ = 4.35 × 10^-4^), subarachnoid haemorrhage (EPVS; HR = 1.53, *P*_FDR_ = 1.22 × 10^-5^), tobacco abuse (EPVS; HR = 1,53, *P*_FDR_ = 1.14 × 10^-11^), cerebral infarction (EPVS; HR = 1.52, *P*_FDR_ = 2.27 × 10^-10^), and TIA (EPVS; HR = 1.51, *P*_FDR_ = 0.001). Additional significant associations were identified for alcohol abuse, speech and voice symptoms, sleep apnoea, malaise and fatigue, polyneuropathy, Parkinson’s disease, epilepsy, depression, and anxiety (HR ranges = 1.13 – 1.47, all *P*_FDR_ < 0.05; Supplementary Table 5).

### Atlas of CSVD neuroimaging biomarker-metabolite associations

Metabolome-wide association analyses were conducted to examine the relationships between circulating metabolic biomarkers and the six CSVD biomarkers (Fig. 5a). In total, 1,365 significant pairs between metabolites and CVSD biomarkers (*P*_FDR_ < 0.05) were identified, with the largest numbers observed for the Fazekas score (n = 263) and CMB (n = 261), followed by PWMH (n = 232), EPVS (n = 218), DWMH (n = 207), and LI (n = 184). The most significant negative associations were found for PUFA_pct with PWMH, DWMH, Razekas score, EPVS, and LI (all *P*_FDR_ < 1 × 10^⁻24^), representing disrupted polyunsaturated fatty acid composition involved in membrane fluidity and anti-inflammatory lipid signaling. CMB was most significantly negatively associated with Omega_6_pct (*P*_FDR_ = 5.17 × 10^⁻34^), reflecting unsaturated fatty acid metabolism and oxidative lipid turnover. Top positive associations include GlycA, a systemic inflammatory marker derived from glycoprotein acetylation, showing strong associations with PWMH (*P*_FDR_ = 5.4 × 10^⁻21^), Fazekas score (*P*_FDR_ = 5.3 × 10^⁻35^), and CMB (*P*_FDR_ = 1.62 × 10^⁻30^). L_HDL_PL_pct, reflecting the phospholipid content of large high-density lipoprotein particles involved in reverse cholesterol transport and endothelial protection, showed most significant associations with DWMH (*P*_FDR_ = 4.61 × 10^⁻20^) and LI (*P*_FDR_ = 8.87 × 10^⁻11^).

Intersections across significant metabolites (Fig. 4b) revealed both shared and feature-specific metabolic associations among the six CSVD inference scores, with 142 metabolites overlapping across all CSVD biomarkers. Other top overlaps were observed among PWMH, DWMH, Fazekas score, EPVS, and CMB (n = 42). Feature-specific patterns were also evident: six metabolites were selectively associated with CMB, whereas three metabolites were selectively associated with LI.

**Fig. 4.**
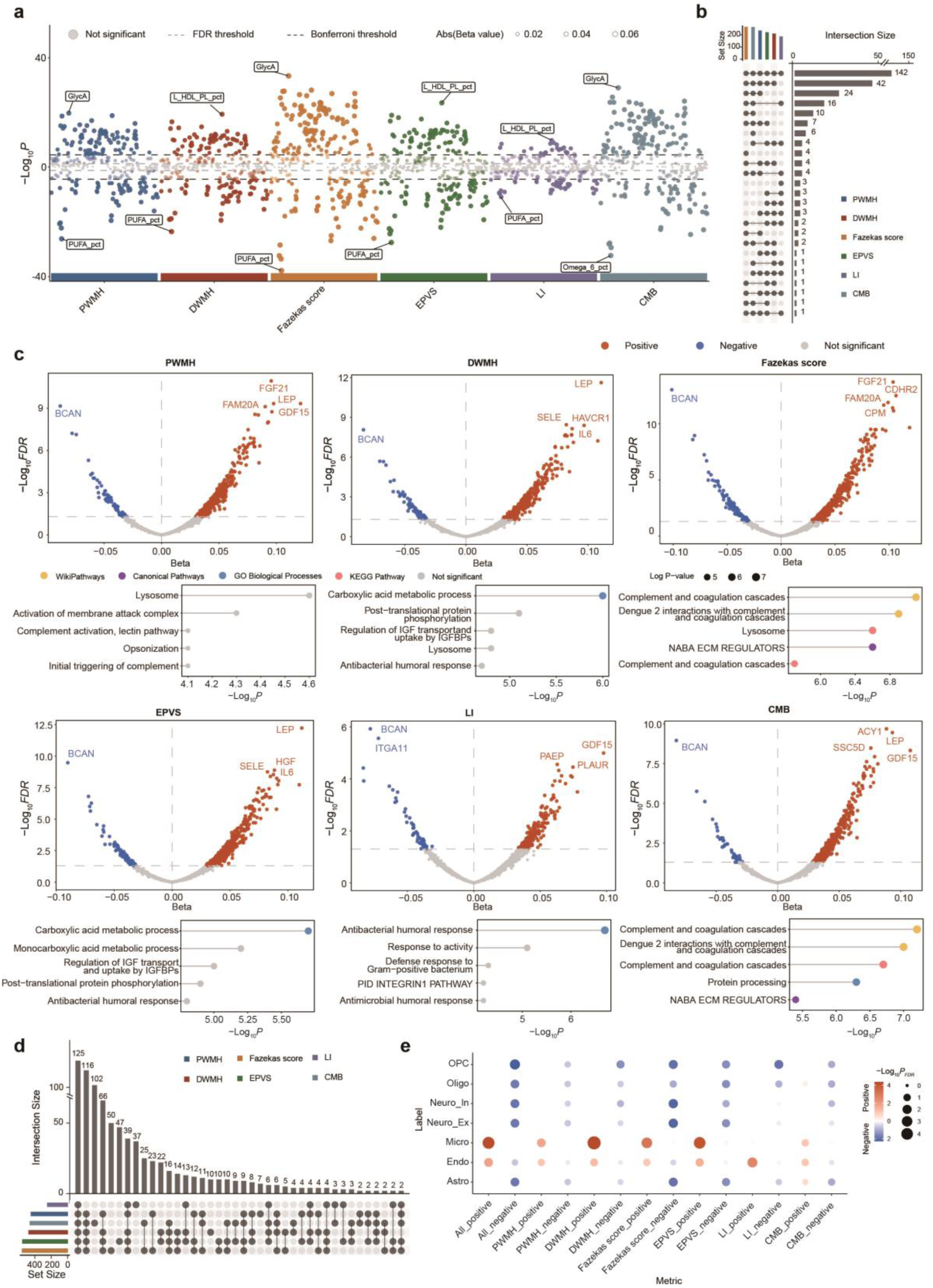
Metabolic and proteomic associations with CSVD biomarkers. (a) Manhattan plot showing results of metabolome-wide association analyses between circulating metabolic biomarkers and six CSVD neuroimaging biomarkers. Each point represents a single metabolite, colored by association direction (red = positive, blue = negative). The dashed line indicates the significance threshold of *P*_FDR_ = 0.05. (b) Upset plots showing shared and biomarker-specific metabolic associations across CSVD biomarkers. Bar heights represent the number of significant metabolites per biomarker, and connected dots indicate overlap between CSVD biomarkers. (c) Volcano plots show biomarker-specific protein associations from proteome-wide analyses. Points are colored by association direction (red = positive, blue = negative; grey = non-significant). Horizontal pathway-enrichment panels beneath each plot show top enriched biological processes. (d) UpSet plot summarizes shared and unique protein associations among CSVD biomarkers. Bars denote the number of proteins per biomarker, and colored lines showed overlapping sets. (e) Heatmap dot plot shows cell-type enrichment of CSVD-associated proteins. Dot size indicates the number of proteins overlapping with the marker genes of each cell type, and color represents the significance of enrichment.

### Atlas of CSVD neuroimaging biomarker-protein associations

Comprehensive proteome-wide association analyses were performed to examine the relationships between 2,920 plasma proteins and the six CSVD biomarkers (Fig. 4). In total, 2,737 significant pairs between proteins and CSVD biomarkers (*P*_FDR_ < 0.05) were identified, with the largest numbers observed for the Fazekas score (n = 556) and EPVS (n = 552), followed by DWMH (n = 475), CMB (n = 461), PWMH (n = 447), and LI (n = 247). Intersections across significant proteins (Fig. 4d) revealed both shared and feature-specific protein associations among the six CSVD biomarkers, with 125 proteins shared across all biomarkers. Notably, we also found unique associations with single CSVD biomarker, including CMB (n = 102), EPVS (n = 47), LI (n = 37), and DPVS (n = 10).

We further explored the heterogeneity of protein associations across the six CSVD biomarkers (Fig. 4c). Across all six CSVD biomarkers, BCAN, involved in synaptic stability and perineuronal net organization, showed most significant negative associations (all *P*_FDR_ < 1 × 10^⁻5^). Distinct positive associations were observed between protein signatures and specific CSVD biomarkers. Specifically, FGF21, involved in regulating lipid oxidation, glucose metabolism, and vascular stress responses, was most significantly associated with PWMH (*P*_FDR_ = 1.13 × 10^⁻11^) and Fazekas score (*P*_FDR_ = 1.91 × 10^⁻14^). LEP, crucial for regulating energy balance and endothelial inflammation, exhibited the most significantly association with DWMH (*P*_FDR_ = 2.4 × 10^⁻12^) and EPVS (*P*_FDR_ = 5.9 × 10^⁻13^). GDF15, essential for mitochondrial stress modulation, inflammation, and vascular remodeling, was most significantly associated with LI (*P*_FDR_ = 1.02 × 10^⁻5^). Lastly, ACY1, involved in amino acid metabolism and neuronal maintenance, was most significantly associated with CMB (*P*_FDR_ = 2.07 × 10^⁻10^).

Pathway enrichment analyses identified biological processes associated with each CSVD biomarkers (Fig. 4c). Gene Ontology (GO) biological process enrichment revealed significant over-representation of proteins in carboxylic acid metabolic process for DWMH (*P*_FDR_ = 1 × 10^⁻6^) and EPVS (*P*_FDR_ = 2 × 10^⁻6^), antibacterial humoral response for LI (*P*_FDR_ = 2× 10^⁻7^), and protein processing for CMB (*P*_FDR_ = 5.01× 10^⁻7^). Additional enrichment was identified across multiple pathway databases, including complement and coagulation cascades and Dengue 2 interactions with complement activation (CMB), extracellular matrix regulation (Fazekas score), lysosomal function (Fazekas score), and complement and coagulation cascades (Fazekas score and CMB). Furthermore, cell-type enrichment analyses demonstrated predominant enrichment of CSVD-associated proteins in endothelial, astrocytic, and oligodendrocytic lineages (Fig. 4e).

### Genetic associations of CSVD neuroimaging biomarkers

Genome-wide association analyses across 7.9 million common variants (MAF ≥ 0.01) in 43,993 UKB European-ancestry participants identified 96 significant variant-trait associations (Supplementary Table 6) and 71 significant locus-trait associations at *P* < 5.0×10^⁻8^/6 = 8.33×10^⁻9^ (Fig. 5a, Supplementary Table 7). When merging association signals and loci of all the six CVSD biomarkers, we found 40 independent association signals and 31 independent loci (Supplementary Table 8 and 9). Specifically, Fazekas score exhibited the largest number of significant associations (n = 26 loci), followed by PWMH (n = 18), EPVS (n = 12), DWMH (n = 12), CMB (n = 2), and LI (n = 1). The most significant genetic association was observed between rs3744017 variant near *TRIM47* and Fazekas score (*P* = 2.73×10^⁻53^, Fig. 5b), with rs55897749 variant near *DCAKD* exhibited a strong association with PWMH (*P* = 5.34×10^⁻22^, Fig. 5c). SNP-based heritability estimates indicated that the common variants explained a moderate proportion of the variance of neuroimaging-based CSVD burden (*P*_FDR_ < 0.05), with the largest heritability observed for Fazekas score (*h*² = 0.28) and PWMH (*h*² = 0.25), followed by EPVS (*h*² = 0.19), DWMH (*h*² = 0.19), CMB (*h*² = 0.15), and LI (*h*² = 0.09). The intercepts of linkage disequilibrium score regression (LDSC) was close to 1 (0.999 to 1.012), which indicated that the slightly elevated lambda (*λ*_*GC*_: 1.05 to 1.14) were more likely attributable to polygenicity rather than to population stratification. Shared genetic architecture analyses revealed overlapping independent variants and genes among CSVD biomarkers, with unique genetic architecture associated with individual biomarkers (Fig. 4d–e). Variant-level signals were most prominent for the Fazekas score (n = 17; Fig. 4d), whereas gene-level overlaps were strongest between the Fazekas score and PWMH (n = 36), followed by gene-level associations unique to the Fazekas score (n = 25; Fig. 4e). Given the high genetic correlation between CSVD neuroimaging traits (*r*_g_ > 0.9, expect for CMB where *r*_g_ > 0.6), we further conducted multi-trait analysis of GWAS (MTAG) to boost statistical power, which could incorporate information across highly genetically correlated traits and adjust for sample overlaps (Supplementary Table 10). As expected, additional variants were discovered, with the highest gain observed for LI (n=30 additional variants), followed by DWMH/EPVS (n=15), CMB (n=13), PWMH (n=8) and Fazekas score (n=1). When merging the loci of all six traits, MTAG discovered two additional loci associated with CSVD neuroimaging biomarkers. A locus located near the *TIE1* gene, where rs3768046 was the lead variant, plays a crucial role in endothelial cell survival and microvascular integrity^18,19^.

**Fig. 5.**
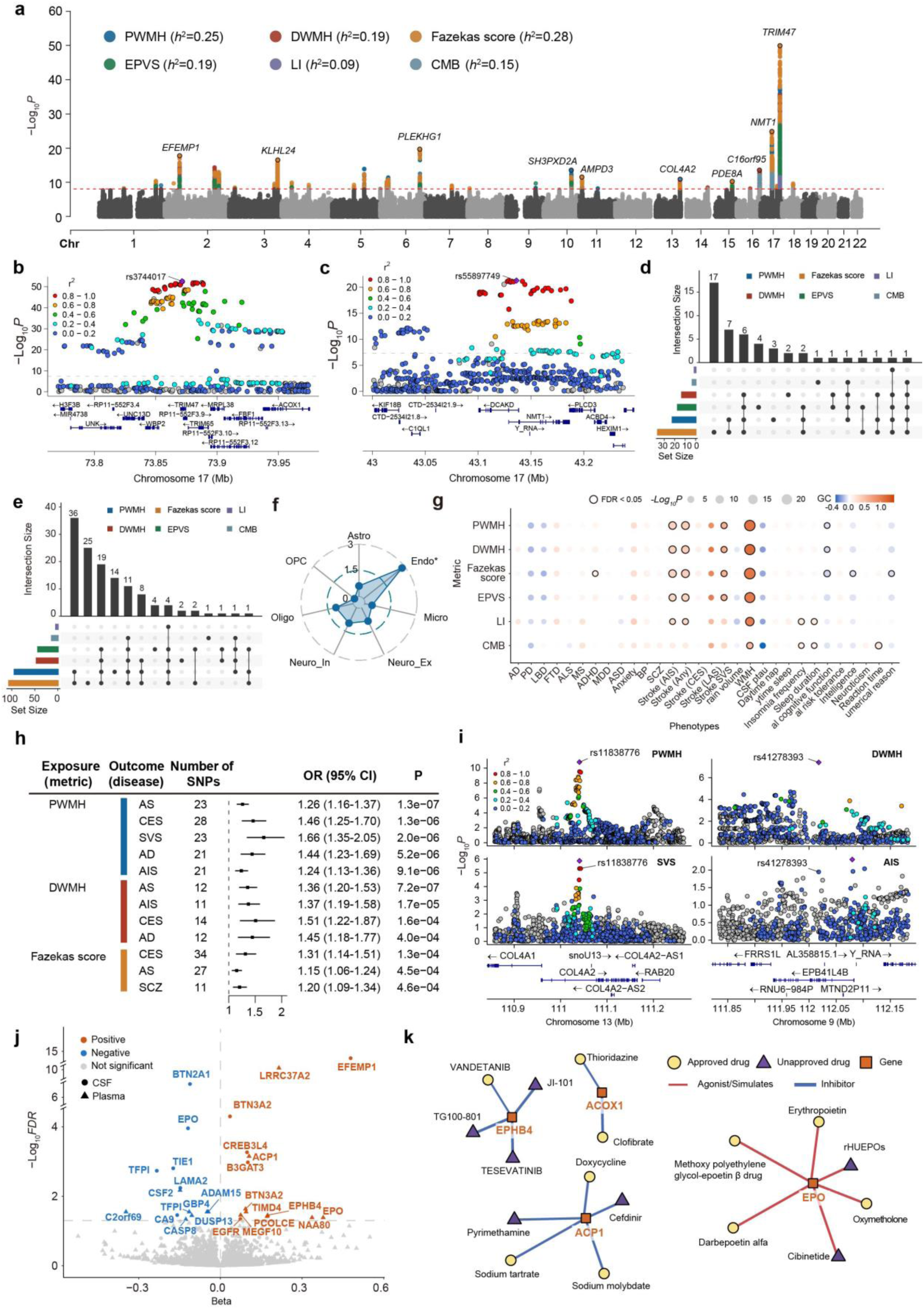
Genetic architecture and therapeutic targeting of CSVD biomarkers. (a) Manhattan plot of genome-wide association analyses for CSVD biomarkers. Each point is a genetic variant colored by the corresponding CSVD neuroimaging biomarker, and the top hits were annotated by the nearest genes. The horizontal red dashed line marks the genome-wide significance threshold (*P* = 5×10⁻⁸/6=8.33×10^⁻9^). Legends represent SNP-based heritability (*h*²). The regional plot of the top hit associated with Fazekas score (b) and PWMH (c), Each point represents a variant colored by linkage disequilibrium (r²) with the lead SNP. (d). UpSet plot of unique and shared genome-wide significant independent variants across CSVD neuroimaging biomarkers. Left bars show per-biomarker set sizes; right bars show intersection sizes. (e) UpSet plot of unique and shared mapped genes from significant loci and gene-based analysis across CSVD neuroimaging biomarkers. (f) Radar plot of cell-type enrichment for genes associated with CSVD biomarkers. X-axis corresponds to major brain cell types (Endo, Micro, Astro, Oligo, OPC, Neuro_Ex, Neuro_In). The polygon radius indicates enrichment ratio. (g) Bubble heatmap of genetic correlations between CSVD biomarkers (rows) and brain-related traits and diseases (columns). Bubble size encodes significance (−Log₁₀*P*); color encodes the genetic correlation estimate (blue to orange scale). Stroked circles denote *P*_FDR_ < 0.05. (h) Forest plot of two-sample MR using CSVD neuroimaging biomarkers as exposures and disease outcomes as outcomes. (i) Regional plots showing the genetic colocalization of PWMH and SVS at rs11838776, as well as DWMH and AIS at rs41278393; (j) Volcano plot of cis-MR between plasma and CSF and CSVD neuroimaging biomarkers. Each point represents a protein. The triangles denote plasma and circles denote CSF, while color indicates the direction of MR relationships (red = positive, blue = negative). The y-axis shows −Log₁₀FDR and the significant proteins with significant cis-MR relationships are labeled. (k) Drug–gene interaction network for proteins supported by cis-MR. Squares represent target genes; yellow circles are approved drugs and purple triangles are investigational drugs. Edge color represents the mode of action of each drug–gene pair. Endo, Endothelial cells; Micro, Microglia; Astro, Astrocytes; Oligo, Oligodendrocytes; OPC, Oligodendrocyte precursor cells; Neuro_Ex, Excitatory neurons; Neuro_In, Inhibitory neuron; MR, Mendelian randomization; AS, Any Stroke; CES, Cardioembolic stroke; SVS, Susceptibility vessel sign; AIS, Acute ischemic stroke; AD, Alzheimer’s disease; SCZ, schizophrenia.

Compared with existing study of CSVD neuroimaging biomarkers, our univariate GWAS summary statistics showed higher heritability than previously reported GWAS summary for EPVS^20^ (*h*² = 0.11), WMH^21^ (*h*² = 0.25) and LI^22^ (*h*² = 0.01). Moreover, we discovered 17, 12, 11, 4, 4 and 1 novel variants associated with Fazekas score, EPVS, PWMH, DWMH, CMB and LI, respectively. Notably, when merging the loci of all six traits, our GWAS identified 14 novel loci associated with CSVD neuroimaging biomarkers. In particular, a locus near the *ARHGAP29* gene, where rs6674226 was the lead variant, regulates signaling and activities in endothelial cells and plays an essential role in blood vessel tubulogenesis^23^. Another locus near the *PLD1* gene, where rs72622537 was the lead variant, is emerging to be involved arterial thrombosis and ischemic stroke^24^ (Supplementary Table 11).

The gene-based association analyses of common variants (MAF ≥ 0.01) were performed using MAGMA, which examined the combined effect of variants in the gene on neuroimaging biomarkers of CSVD. A total of 258 gene-wide significant associations were observed at *P* < 0.05/18,879/6 = 4.41×10^⁻7^ (Supplementary Table 12), with the highest number of associations observed for PWMH (n = 85) and Fazekas score (n = 84), and the least observed for LI (n = 4). Of note, *TRIM65*^25^, *COL4A2*^26^, and NMT1^27^, which are well-known to be associated with CSVD neuroimaging traits, were significant in both univariate GWAS and gene-based association analysis. In contrast, *TFPI*^28^, *GFAP*, *APOC1* and *HTRA1*^29^, which are associated with the pathogenesis and severity of CVSD, were only discovered in the gene-based association analysis. Moreover, the gene-based burden test of rare variants (MAF < 0.01) conducted with SAIGE-GENE+ did not identify significant associations at Bonferroni-correlated *P* < 0.05/18,731/6 = 4.45×10^⁻7^.

The function of lead variants of the loci identified by univariate GWASs and MTAG were annotated using Ensembl Variant Effect Predictor (VEP) and mapped to 41 unique genes. the 60 unique lead variants of the discovered loci were predominantly located in intronic region (61.67%), followed by intergenic region (8.33%) and upstream region (8.33%). In particular, the rs11838776 variant located in intronic region of *COL4A2* gene has been associated with both white matter hyperintensities and lacunar stroke, suggesting a shared genetic contribution to CSVD-related microvascular injury^22^. Additionally, the rs3744017 variant located in intronic region of *TRIM47* has been associated with increased white matter hyperintensity burden in European populations^30^. We pooled these genes with those identified by gene-based analysis to conduct cell-type enrichment analyses and examine potential functional enrichment. We found that CSVD-associated genes were exclusively and significantly positively enriched in endothelial lineages (*P*_FDR_= 0.005, Fig. 5f), suggesting a predominant vascular involvement in small-vessel pathology.

### Genetic correlations with health-related traits and brain disorders

To explore the genetic correlation between CSVD biomarkers and brain-related traits and disorders, we conducted cross-trait LDSC analyses between six CSVD biomarkers and 13 brain-related traits and 17 major neuropsychiatric disorders (Fig. 5g; Supplementary Table 13). We identified 31 significant associations between CSVD biomarkers and various phenotypes (*P*_FDR_ < 0.05). Notably, PWMH, DWMH, Fazekas score, and EPVS showed significant positive associations with any type of stroke and cardioembolic stroke. In the cognitive domain, greater Fazekas score was genetically correlated with worse general cognitive function (*r*_g_ = −0.10, *P*_FDR_ = 0.001), suggesting that white matter hyperintensities may be linked to cognitive decline. Conversely, CMB showed a positive association with reaction time (*r*_g_ = 0.17, *P*_FDR_ = 0.001), suggesting that microvascular pathology may be related to processing speed. Notably, these results are aligned with our PheWAS findings (Fig.3a), which also highlighted cognitive and processing speed deficits associated with microvascular damage.

### Causal relationships between CSVD neuroimaging biomarkers and brain disorders

To examine potential causal relationships between CSVD neuroimaging biomarkers and clinical outcomes, we performed bidirectional two-sample Mendelian randomization (MR) analyses between six CSVD biomarkers and 17 major neuropsychiatric brain disorders. The forward MR analyses, with CSVD neuroimaging biomarkers as exposure and brain disorders as outcome, identified 12 significant causal effects of three CSVD biomarkers on six neurological and psychiatric disorders (*P* < 0.05/17/6 = 4.9×10^⁻4^, Fig.5h, Supplementary Table 14). Specifically, The positive causal effects of PWMH were strongest for susceptibility vessel sign (IVW OR = 1.66, 95% CI = 1.35 to 2.05, *P* = 2.0×10^⁻6^), followed by cardioembolic stroke (IVW OR = 1.46, 95% CI = 1.25 to 1.70, *P* = 1.3×10^⁻6^), AD (OR = 1.44, 95% CI = 1.23 to 1.69, *P* = 5.2×10^⁻6^), any stroke (IVW OR = 1.26, 95% CI = 1.16 to 1.37, *P* = 1.3×10^⁻7^) and acute ischemic stroke (IVW OR = 1.24, 95% CI = 1.13 to 1.36, *P* = 9.1×10^⁻6^). Similarly, DWMH was causally associated with increased risk of stroke (any type, acute ischaemic, and cardioembolic) and AD. Lastly, Fazekas score was significantly associated with stroke (any type and cardioembolic) and schizophrenia. Sensitivity analysis further confirmed that the identified MR relationships showed no evidence of directional pleiotropy (*P*_BONF_ of all MR-Egger intercept > 0.05) and horizontal pleiotropy (*P*_BONF_ of all MR-PRESSO > 0.05). Reverse MR utilized brain disorders as exposure and CSVD neuroimaging biomarkers as outcome did not identify any significant causal relationships (*P* > 4.9×10^⁻4^, Supplementary Table 15), which further confirmed the CSVD biomarkers were more likely to be a cause rather than a consequence of the brain disorders. All these findings highlight that CSVD biomarkers contribute to the risk of cerebrovascular, neurodegenerative, and psychiatric conditions.

The genetic colocalization of the identified forward MR relationships were further examined using coloc R package. We found that the 12 significant MR relationships also shared the same causal variant at a posterior possibility PP.H4 > 0.80. Specifically, PWMH and usceptibility vessel sign were colocalized at a *COL4A2,* with rs11838776 as the causal variant (PP.H4 = 0.80), while DWMH and acute ischemic stroke were colocalized at a *EPB41L4B* with rs41278393 as the causal variant (PP.H4 = 0.99).

### Drug target identification

To examine whether the CSVD biomarkers with a significant causal effect on brain disorders could inform drug development, we next performed forward *cis*-MR analyses using *cis* pQTL of 2,920 plasma and 7,028 CSF protein levels as exposures and three CSVD biomarkers (PWMH, DWMH, Fazekas score) as outcomes, and reverse MR analysis using the same CSVD biomarkers as exposures and protein levels as outcomes. After excluding reverse MR relationships, a total of 30 and 29 forward *cis*-MR relationships were found for 17 plasma and 12 CSF proteins, respectively (Fig. 5j; Supplementary Table 16). Notably, BTN3A2, ACOX1, EPO and TFPI proteins were found to have significant causal effect on CSVD biomarkers in both CSF and plasma. The most significant positive relationships were observed for EFEMP1, LRRC37A2, BTN3A2, and ACP1, whereas the most significant negative effects were identified for BTN2A1, EPO, TIE1, and TFPI. Functionally, the positive associations were linked to endothelial signalling, angiogenesis, and extracellular-matrix remodelling, which contribute to vascular health and could exacerbate CSVD pathology. The negative associations were tied to thrombosis and inflammation, suggesting impaired vascular function. In sensitivity analysis, we did not observe significant horizontal and directional pleiotropy for the significant *cis*-MR relationships. We also conducted cis-MR analysis for all the six CSVD biomarkers, an additional 11 and 6 *cis*-MR relationships involving 10 and 5 proteins for CSF and plasma proteins were observed (*P*_FDR_ < 0.05, Supplementary Table 17). Together, these findings highlight the convergent vascular mechanisms underlying CSVD and suggest potential therapeutic targets related to endothelial function and inflammation.

By pooling the discovered CSF and plasma proteins together, we further employed systemic search to map significant causal genes to known drug–target interactions that align with the desired modes of action, aiming to identify pharmacological mechanisms that could be leveraged for CSVD-related therapeutic pathways. Of the 25 unique proteins that exhibited significant cis-MR relationships, a total of seven drugs act as agonists/simulates of two proteins (EPO and BTN2A1) and 15 drugs acting as inhibitors of four proteins (EPHB4, ACP1, EGFR and ACOX1) were discovered (Supplementary Table 18). Specifically, one approved (VANDETANIB) and three investigational drugs (TESEVATINIB, JI-101 and TG100-801) were identified as inhibitors of EPHB4 protein. Two approved drugs (Thioridazine and Clofibrate) were identified as annotated inhibitors of ACOX1, a lipid-metabolism enzyme linked to white-matter integrity^31^. In contrast, several approved and investigational EPO-pathway agonists were identified in our drug–target analysis, including Erythropoietin, Darbepoetin alfa, Methoxy polyethylene glycol-epoetin β drug and Oxymetholone, as well as two investigational drugs (rHUEPOs and Cibinetide; Fig.5k), representing upstream activators of the EPO-associated causal protein.

## Discussion

In this study, we present a transformer-based deep learning framework that generates six continuous inference scores representing cerebrovascular small-vessel burden from routine MRI scans. These AI-derived phenotypes capture inter-individual variability in CSVD-related biomarkers beyond conventional visual ratings, revealing a continuous spectrum of microvascular pathology across individuals. Importantly, the robustness and clinical relevance of the model were demonstrated through two independent external validations. In addition to achieving consistent claissification accuracy across external datasets, we show that AI-augmented clinical assessments significantly improve diagnostic accuracy among junior clinicians, underscoring the model’s potential as a decision-support tool in real-world settings. The observed associations between CSVD biomarkers and phenotypic, clinical, and biological signatures highlight this model as a quantitative tool for mapping cerebrovascular health, bridging subclinical microvascular pathology with clinically manifest disease and functional impairments.

The principal advancement of CSVDtransformer that using the novel feature-reranking approach over conventional deep learning architectures like ResNet, ViT, and SWIN transformer lies in its fundamental shift from a spatially-prioritized to a functionally-prioritized model induction, a shift that is particularly critical for capturing the complex and diffuse nature of CSVD biomarkers. Conventional deep learning models are inherently constrained by their inductive bias towards local spatial coherence, using convolutional kernels that assume the most relevant features reside in proximate voxels. Similarly, the standard ViT begins by segmenting the image into rigid, spatially-contiguous patches. This spatial constraint is a profound limitation for CSVD, whose hallmark biomarkers, including widely distributed white matter hyperintensities, subtle lacunar infarcts, cerebral microbleeds, and atrophy, are often spatially dispersed throughout the brain’s subcortical and deep white matter regions, forming a collective burden rather than a localized lesion. Our method directly counteracts this by leveraging the foundational pre-processing step of spatial registration to perform a voxel-wise statistical screening, ranking and grouping voxels based on the mean of absolute value of their correlation with CSVD biomarkers. This forms patches that are functionally coherent with the disease’s pathological signature, meaning a single patch could intelligently combine a voxel from a periventricular white matter lesion with another from a strategically located lacune, precisely because they share a similar statistical relationship to the clinical label. This creates a powerful, CSVD-specific inductive bias from the outset, ensuring that the model’s computational budget, especially the transformer’s self-attention mechanism, is focused exclusively on assembling these distributed, multi-faceted biomarkers into a holistic disease score. This leads to a more data-efficient and robust learning process, as the model is not forced to rediscover these diffuse patterns from scratch, and it profoundly enhances clinical interpretability. The initial feature ranking provides a direct, univariate map of voxels most implicated in CSVD, while the subsequent transformer attention can reveal how interactions between different types of lesions (e.g., the co-occurrence of microbleeds and white matter damage) contribute to the overall clinical picture, ultimately offering a more sensitive and nuanced tool for quantifying CSVD burden than methods anchored in spatial locality alone.

A major challenge in translating AI models has been their limited generalizability across populations, imaging protocols, and clinical settings. In practice, AI performance often degrades when applied to independent cohorts with heterogeneous imaging protocols and population characteristics, underscoring the importance of robust external validation. Here, we addressed this challenge through two complementary external validation strategies. First, the model exhibited stable classification accuracy across independent external datasets, supporting its robustness beyond the training distribution. Second, we assessed the model under a realistic diagnostic scenario by examining whether access to AI-argumented assessments could improve the performance of junior clinicians. The observed increase in diagnostic accuracy indicates that the model provides clinically meaningful information that can support decision-making when domain expertise is limited. Together, these findings suggests that CSVDTransformer does not merely replicate expert-level performance, but also practically useful, with potential to standardize CSVD assessment and reduce variability in diagnostic performance across different levels of clinical experience.

Leveraging these robust and generalizable inference scores, we next examined their clinical and biological relevance across large-scale population datasets. Across phenome-wide analyses, higher inference score of CSVD biomarker was associated with increased risks of neurodegenerative and psychiatric disorders, extending the impact of small-vessel pathology beyond classical vascular outcomes. These findings reveal a transdiagnostic pattern of cerebrovascular vulnerability, resonating with prior reports by demonstrating that CSVD burden extends beyond vascular and cognitive outcomes to encompass a wide range of affective, psychotic, and behavioral manifestations^32^. Notably, CSVD biomarkers showed significant associations with sleep, personality, mental health, and cognitive traits, highlighting the broad behavioral and neuropsychiatric footprint of microvascular burden. Such multi-domain associations support the notion that cerebrovascular pathology influences brain health through partially dissociable but overlapping vascular–affective–cognitive pathways. Past findings have suggested that white matter hyperintensities broadly affecting global cognition, executive function, memory, and attention, while cerebral microbleeds are more strongly linked to executive and attentional deficits, and lacunes predict decline in executive function and psychomotor speed, supporting the notion of partially dissociable vascular–cognitive mechanisms^33^. Collectively, these results position cerebrovascular burden as a core dimension of brain health, shaping vulnerability across behavioral, affective, and cognitive domains and offering a vascular framework for understanding transdiagnostic brain disorders.

Integrative multi-omic analyses revealed that the CSVD biomarkers converge on pathways involved in lipid metabolism, endothelial integrity, immune–inflammatory signalling, and systemic homeostasis. Metabolic and proteomic analyses highlighted enrichment in lipid metabolic and complement activation pathways, implicating endothelial inflammation and immune–metabolic dysregulation in cerebrovascular vulnerability. Several of the top protein correlates, including *SELE*, *HGF*, *BCAN*, and *PLAU*, are key regulators of endothelial activation and extracellular matrix re-modeling, processes central to blood–brain barrier integrity. These pathways converge on systemic metabolic–vascular and immune–endothelial interactions that are increasingly recognized as contributors to microvascular integrity and CSVD burden^34,35^. Genetic associations converged on loci such as *COL4A1* and *COL4A2*, which encode basement membrane proteins critical for endothelial integrity and vascular stability^36,37^, aligning with prior evidence linking extracellular matrix dysfunction to small-vessel pathology and stroke risk. Moreover, drug-target analyses identified plausible pharmacological agents acting on ACOX1, EPO, and EPHB4, highlighting convergent pathways related to lipid metabolism^31,38^, neurovascular repair^39^, and vascular morphogenesis^40^, respectively. These findings suggest that pharmacological modulation of these CSVD-relevant biological pathways may offer avenues for therapeutic repurposing. However, our preliminary results should be viewed as hypothesis-generating, given that the identified agents were not designed for CSVD, and their relevance to CSVD pathology remains untested. Future work is needed to establish whether modulation of these protein pathways has translational relevance.

As the first study incorporating various cerebrovascular imaging markers at the population level, our proposed AI model offer a scalable, objective, and reproducible digital phenotype of cerebrovascular health. These indices can facilitate large-scale disease monitoring and early risk stratification, demonstrating its potential for widespread clinical and epidemiological applications. In clinical contexts, such quantitative measures could assist patient selection and treatment monitoring in trials targeting vascular and neurodegenerative diseases, by capturing subtle cerebrovascular changes that may precede overt clinical symptoms. More broadly, integrating AI-derived CSVD scores into multi-omic and longitudinal frameworks could accelerate precision prevention strategies and individualized prognostic modeling. The observed associations between CSVD burden and diverse brain disorders further underscore the clinical relevance of incorporating cerebrovascular health metrics into early detection and intervention pipelines.

Several methodological constrains warrant discussion. First, our model was developed using UK Biobank data, a predominantly community-based cohort that underrepresents individuals with neurological or cerebrovascular diseases. This limitation may reduce the representation of participants with clinically significant or symptomatic brain disorders, potentially constraining the model’s generalizability to patient populations. External validation in multi-ethnic and multi-center clinical cohorts is therefore warranted to ensure robustness across diverse disease spectra. As the present analyses are primarily cross-sectional, longitudinal imaging and clinical follow-up are required to capture temporal dynamics and clarify causal relationships between cerebrovascular burden and disease progression. Finally, the current model was trained on T2-FLAIR and SWI scans, which primarily reflect white matter and microbleed pathology. Future training model incorporating additional modalities, such as perfusion imaging, retinal microvasculature, and systemic biomarkers, may help build a more integrated framework linking brain, vascular, and peripheral systems.

Our transformer-based model provides a scalable framework for quantifying cerebrovascular small-vessel burden from routine MRI, offering a reproducible and interpretable digital marker of vascular brain health. By enabling continuous assessment of CSVD burden, this approach bridges clinical imaging with population-scale and biological analyses, facilitating early detection, individualized risk profiling, and integration into precision prevention pipelines. The observed associations between AI-inferred CSVD scores and diverse brain disorders underscore the model’s potential to uncover shared vascular mechanisms underlying neurological and psychiatric diseases. Collectively, these advances move cerebrovascular research toward a proactive, data-driven paradigm for early intervention and personalized brain health monitoring.

## Methods

### UK Biobank study population

The UK Biobank included phenotypic and genetic information for over 500,000 participants of ages between 40 and 69 across the UK^41^. Informed consent has been signed by all participants. The UK Biobank cohort was approved by the NHS National Research Ethics Service North West (reference number: 16/NW/0274). The data utilized in the study included neuroimaging data (T1, T2-FLAIR and SWI), diagnostic information, demographic data, brain health related traits, genetic profiles, and plasma proteomic data. The research was performed under application number 202239 and 19542.

### External datasets and MRI acquisition

Two independent external validation cohorts were recruited from Huashan Hospital, Shanghai, China (Huashan dataset 1, N = 176; Huashan dataset 2, N = 392), as part of the Vascular, Imaging and Cognition Association of China (VICA) study^42^. These cohorts were completely independent from the training and internal validation datasets, and no participants from the external cohorts were involved in model development, hyperparameter tuning, or threshold selection.

All participants underwent standardized brain MRI examinations, including T1-weighted imaging, T2-weighted fluid-attenuated inversion recovery (T2-FLAIR), and susceptibility-weighted imaging (SWI). These multimodal MRI data were used to evaluate the generalizability of the CSVDTransformer model. MRI data were acquired on 3.0-Tesla MRI scanners (GE, Siemens) using standardized clinical protocols. T1-weighted images were acquired using a three-dimensional T1-weighted sequence with repetition time (TR) = 1,450 ms, echo time (TE) = 25 ms, and slice thickness = 4 mm, flip angle = 111°. T2-FLAIR images were acquired with TR = 8,000 ms, TE = 113.5 ms, flip angle = 160°, and slice thickness = 2 mm. SWI data were acquired using a three-dimensional gradient-echo sequence with TR = 54 ms, TE = 23.5 ms, flip angle = 15°, and slice thickness = 2 mm.

### Neuroimaging CSVD biomarkers

Six CSVD imaging biomarkers were included in the present study. Among them, periventricular white matter hyperintensities (PWMH), deep white matter hyperintensities (DWMH), Fazekas score, enlarged perivascular spaces (EPVS), and lacunes were derived from 3,718 T2-FLAIR scans, whereas CMB was derived from 2,774 SWI scans. All imaging data were pre-processed and quality-controlled following the UK Biobank imaging protocol, and visual ratings were performed by trained raters according to established criteria.

All neuroimaging markers of cerebral small vessel disease were evaluated according to the STRIVE criteria. Eight physicians with 6 to 10 years of clinical experience independently annotated the images and reached consensus through joint review in cases of disagreement. White matter hyperintensities (WMHs) of presumed vascular origin were defined as hyperintense lesions on T2-weighted and FLAIR images in the periventricular or deep white matter without cavitation, following STRIVE recommendations. The severity of WMHs was rated using the Fazekas scale, where periventricular WMH was graded as 0 (absent), 1 (caps or pencil-thin lining), 2 (smooth halo), and 3 (irregular hyperintensities extending into the deep white matter), and deep WMH as 0 (absent), 1 (punctate), 2 (beginning confluence of foci), and 3 (large confluent areas). Fazekas score was calculated by summing both periventricular and deep scores, thus had a level of 0 to 6. Lacunes of presumed vascular origin were defined as round or ovoid, subcortical, fluid-filled cavities with cerebrospinal fluid–like signal intensity (3–15 mm in diameter) on all sequences, sometimes with a thin hyperintense rim on FLAIR, located in typical perforating artery territories; enlarged perivascular spaces (<3 mm) and cortical infarcts were excluded. Enlarged perivascular spaces (EPVS) were defined according to the STRIVE criteria as small (<3 mm), round or linear CSF-like signal intensities following the course of penetrating vessels, without a FLAIR hyperintense rim. EPVS burden was rated visually in the basal ganglia on one side of the brain using a three-category scale (category 1 < 10, category 2 = 10–25, category 3 > 25 visible EPVS), as previously described by Doubal et al.^2^. Cerebral microbleeds were identified as small (<10 mm), round or ovoid hypointense lesions on T2*-weighted or susceptibility-weighted imaging, consistent with hemosiderin deposits, excluding vascular flow voids, calcifications, and artifacts. To assess inter-rater consistency, 10% of the images were re-evaluated by two independent raters. For downstream modeling, EPVS, lacunes, and CMB were binarized to indicate the presence or absence of each marker in a given subject.

### Neuroimaging data preprocessing

The UK Biobank brain imaging were preprocessed using FSL and Freesurfer, following their official pipelines^43^. The main steps begin with the defaced, raw full field-of-view (FoV) image. This image first underwent a reduction of its FoV to minimize non-brain tissue, followed by Gradient Distortion Correction, a process robustly achieved using FSL-BET^44^ and FSL-FLIRT^45^ in conjunction with an MNI152 template to produce the reduced-FoV T1 head image. Next, the data was nonlinearly warped to 1 mm^3^ MNI152 standard space using FSL-FNIRT. The T2-FLAIR and SWI were linearly aligned to the T1 using FLIRT, and the resulting warp were combined to resample T2 FLAIR and SWI into 1 mm^3^ MNI152 standard space. Subcortical and white matter regions were extracted for subsequent model analysis. All external MRI data from Huashan datasets were preprocessed using the same standardized pipeline as the UK Biobank dataset, without any additional fine-tuning or recalibration of the model on the external cohorts, ensuring a strict and unbiased assessment of model generalizability.

### CSVDtransformer model

The CSVDtransformer architecture introduces a novel feature-reranking mechanism to optimize the identification of complex CSVD patterns from multi-modal structural MRI (T1w, T2-FLAIR, SWI). The model begins by leveraging the spatial normalization of all brain images to a common template, a prerequisite that enables a voxel-wise statistical pre-screening. Specifically, we computed the absolute correlation coefficient between the intensity of each voxel across the training cohort and the target CSVD biomarker labels. This process generates a whole-brain correlation map that ranks all voxels based on their individual discriminative power. Instead of using conventional spatially contiguous patches, we then vectorized the brain image and form our initial input tokens by grouping the voxels into functionally coherent, non-spatially contiguous patches based on their average absolute correlation with the labels. These patches, which may intelligently aggregate biomarkers from disparate brain regions that collectively signal CSVD biomarkers, are then projected into a latent embedding space. This embedding, further with positional information, is subsequently processed by a standard transformer encoder. The encoder’s self-attention mechanism then learns the complex, multivariate interactions between these highly salient, pre-selected patches to generate a final prediction for the CSVD biomarker. This end-to-end framework ensures that the model’s representational capacity is focused from the outset on the most informative biomarkers, providing a powerful inductive bias for capturing the diffuse and multi-focal nature of CSVD pathology.

The CSVDtransformer was trained on a randomly selected cohort of 3,000 subjects. The remaining data were divided into internal validation and testing sets. The validation set was used for hyperparameter tuning, which included the number of layers^46^, patch size [512, 1024, 2048], and embedding dimensions [128, 256, 512]. The model was trained for 50 epochs with a batch size of 256 using the AdamW optimizer. A cosine learning rate scheduler with 5 epochs of warm-up was employed, with an initial learning rate of 1e-4. For the prediction of CMB, model inputs were SWI and T1-weighted images. For the prediction of the other five biomarkers, inputs were T2 FLAIR and T1-weighted images. The model was trained for simultaneous multi-biomarker prediction using a multi-class classification cross-entropy loss. The contributions of the different loss functions were balanced^47^. For comparison, baseline models, including ResNets, Vision Transformer (ViT), and SWIN transformers, were trained on the same dataset. Each baseline model was trained with its own set of optimized hyperparameters.

### Exposure variables

Six AI-derived cerebrovascular inference scores corresponding to the above imaging biomarkers (PWMH, DWMH, Fazekas score, EPVS, lacunes, and CMB) were used as exposure variables in subsequent analyses. Each score represented the model-inferred probability or burden of the respective CSVD feature and was standardized (z-score) within the analysis sample prior to statistical modeling.

### Phenome-wide association analysis

We performed PheWAS to examine the association of CSVD inference scores with brain disorders and brain health traits. Traits-related analyses covered 4 categories (cognition, mental health and well-being, sleep, personalities) and 98 specific traits (Supplementary Table 1). The disease-related analyses initially considered four categories and 138 specific diseases (Supplementary Table 3). Disease diagnoses were ascertained from hospital inpatient records, primary care data, and death registries, with follow-up censored at the last follow-up record^48^. To ensure reliable estimation, only diseases with ≥50 prevalent cases for cross-sectional analyses or ≥50 incident cases for longitudinal analyses were retained for modeling.

### Associations with brain health-related traits

We examined the associations between CSVD inference scores and a comprehensive set of brain health–related traits derived from the UK Biobank (UKB) baseline assessments. Following our prior research^49^, we included 98 brain health factors covering four domains: cognition (n = 12), mental health and well-being (n = 65), sleep (n = 6), and personalities^50^ (n = 15).

Associations between each CSVD inference score and brain health trait were tested using linear regression models adjusted for age, sex, imaging site, total intracranial volume, education, body mass index (BMI), years of education, and Townsend deprivation index. All continuous variables were standardized to z-scores before analysis. Multiple testing correction across all traits and CSVD biomarkers was performed using the Benjamini–Hochberg false discovery rate (FDR) method, with p < 0.05 considered statistically significant. Detailed variable definitions, coding schemes, and corresponding UKB field IDs are provided in Supplementary Table 1.

### Cross-sectional analysis of disease phenotypes

Cross-sectional associations between each imaging phenotype and disease prevalence were evaluated using multivariable logistic regression models. For each endpoint, prevalent cases were defined as those with diagnosis dates preceding the imaging visit, while participants without any diagnosis of that disease served as controls. Participants who developed the disease after MRI were excluded from cross-sectional analyses. Models were adjusted for key demographic and imaging covariates, including age at scan, sex, head size, assessment center, and additional modality-specific confounders. Odds ratios (ORs) and 95% confidence intervals (CIs) were estimated for each exposure–disease pair, and false discovery rate (FDR) correction was applied across outcomes.

### Longitudinal risk analysis of disease outcome

For prospective outcomes, we employed Cox proportional hazards regression to estimate hazard ratios (HRs) of incident neurological and psychiatric disorders associated with baseline CSVD burden. Disease endpoints were defined according to ICD-10 codes (Supplementary Data 1) and derived from the UK Biobank “first occurrence” data (categories 2401–2417), which integrate records from primary care (category 3000), hospital inpatient (category 2000), self-reported medical conditions (field 20002), and national death registries (fields 40001 and 40002). Follow-up starts from the initial UK Biobank assessment visit and ends at the earliest of disease diagnosis, death, or the last available hospital or primary care record, whichever occurred first. For each endpoint, individuals with self-reported diagnoses or with any record of the corresponding disease category prior to baseline were excluded. Proportional hazards assumptions were checked using Schoenfeld residuals. Age, sex, and scanner site were controlled as covariates of non-interest.

### Metabolic and protein association analysis

Baseline plasma samples from the UK Biobank were processed using the Olink Explore 3072 platform on a NovaSeq 6000 Sequencing System, which quantified the relative abundance of 2,923 unique proteins expressed as normalized protein expression (NPX) values ^51,52^. Proteins with a missing rate ≥25% (n = 3; GLIPR1, NPM1, and PCOLCE) were excluded from further analyses. All protein concentrations were inverse-rank normalized, and the effects of chronological age and sex were regressed out before association testing. The plasma samples were also prepared and processed to quantify blood biochemistry, blood count, and metabolic biomarkers^53^. Associations between each CSVD neuroimaging trait and plasma protein levels were examined using a generalized linear model in R. Each model included the following covariates: chronological age at imaging, sex, total brain volume, imaging site, years of education, Townsend deprivation index, smoking status, drinking status, and ethnicity. The significance threshold was set at FDR-corrected *P* < 0.05 across all 2,920 × 6 protein–CSVD biomarker pairs using the Benjamini–Hochberg procedure. Functional enrichment analysis of significant proteins was performed using over-representation analysis in Metascape (https://metascape.org/), and cell type enrichment was evaluated using over-representation analysis from Python SciPy^54^, with the maker genes of seven brain cell types obtained from previous studies^54,55^ and all coding genes of 2,920 proteins utilized as background gene set. Similar generalized linear models were performed for blood biochemistry, blood count, and metabolic biomarkers, with age at baseline, sex, fasting time and statin used as covariates. The significance was determined by FDR-corrected *P* < 0.05 across all 313 × 6 pairs of blood biomarker and CSVD neuroimaging traits.

### Genotyping and quality control

Genetic data for this study were processed and quality-controlled according to the standard UK Biobank pipeline. All genetic analyses were conducted using GRCh37 genomic coordinates. Genotyping, imputation, and quality control procedures were centrally managed by the UK Biobank team^56^. UKB participants were genotyped using either the UKB Axiom Array or the UK BiLEVE Axiom Array. Following official processing, we further applied additional quality control at both sample and variant level using PLINK v.2.0. At the sample level, we excluded participants with a missing rate > 5%, sex mismatches, abnormal sex chromosome aneuploidy, heterozygosity rate outliers, and those that were not in the white British ancestry subset or had more than ten putative third-degree relatives. At the variant level, variants were excluded if they had a call rate < 95%, imputation quality score < 0.5, minor allele frequency (MAF) < 0.01, or deviated from Hardy-Weinberg equilibrium (*P* < 10⁻⁶). Following these QC steps, a final dataset consisting of 43,993 individuals and 7.9 million variants remained for downstream analyses.

### Genetic association analysis

Univariate GWAS were first conducted for the six CSVD neuroimaging traits using PLINK 2.0. To further boost the power of detecting CSVD neuroimaging trait-associated loci, MTAG was used to conduct multi-trait GWAS, which performs a meta-analysis of GWAS summary for highly correlated traits while allowing for sample overlaps. For all these associations analysis, the effect of age, age², sex, total brain volume, imaging center, brain position in scanner and the first forty genetic principal components (PCs) were regressed out. The significance threshold of univariate GWAS was set to Bonferroni-corrected *P* < 8.33×10^⁻9^ (5×10^⁻8^/6).

The gene-based common-variant association analysis was conducted using MAGMA software for 18,879 protein-coding genes. The significance threshold was set to Bonferroni-corrected *P*< 0.05/18,879/6 = 4.41×10^⁻7^ for the univariate GWAS. Meanwhile, the gene-based rare-variant association analysis was performed using Burden test, SKAT and SKAT-O in SAIGE-GENE+, with age, age², sex, total brain volume, imaging center, brain position in scanner and the first ten genetic PCs included as covariates. The significance was determined at Bonferroni-correlated *P* < 0.05/18,731/6 = 4.45×10^⁻7^.

Based on EUR-specific LDs, we identified LD-independent variant-trait associations using PLINK clumping for each GWAS, with a clumping distance of 3Mb and LD threshold of *r*^2^>0.1. The identified lead variants were defined as independent variants, and the genetic association of these variants were defined as LD-independent variant-trait associations. Furthermore, for each trait, we identified LD-independent locus-trait association based on the following criteria: (1) adding 500kb to both sides of each LD-independent variant to create initial loci; (2) merging overlapping loci using GenomicRanges R package; (3) merge loci if one of the lead variants of one loci was in LD with any lead; (4) all loci overlapping with MHC region and chromosome 8p23.1 were merged into on locus. The remaining loci of the trait were defined as LD-independent locus-trait associations. By pooling locus-trait associations of all CSVD biomarkers, the non-overlapping loci at *r*^2^<0.1were further defined as LD-independent loci. The locus was named by the nearest gene^57,58^. In terms of the definition of novelty, we curated previously reported GWAS summary statistics of the WMH^59^, CMB^60^, LI^22^ and EPVS^20^. A variant-biomarker association was considered as novel when the variant was > 500kb away from and not in LD (*r*^2^>0.1) with any known variants (*P* < 5.0×10^⁻8^) of the same CSVD neuroimaging trait. Similar, we considered an association signal as novel when the variant was > 500kb from and not in LD with any known variants for all the CSVD neuroimaging traits. Finally, a locus was considered novel when all lead variants in the locus were > 500kb away from and not in LD with any known variants for all six CSVD neuroimaging traits^61^.

### Functional annotation

The functional annotation of variants identified with univariate GWAS and MTAG were performed using Ensembl VEP tool. The variants were categorized based on genomic location and functional consequence and mapped to genes. Similar to that in proteomic analysis above, we performed cell type enrichment using over-representation analysis from Python SciPy^54^. The mapped gene from significant variant-based analysis and the unique genes from significant gene-based analysis were pooled together and all the protein-coding genes were used as background. The significance was determined at a FDR-corrected *P* < 0.05.

### Heritability estimation

We estimated the heritability for all six CSVD neuroimaging traits, including both common and rare variants. The common-variant-based heritability was calculated with LDSC software, with age, age², sex, total brain volume, imaging center, brain position in scanner and the first forty genetic principal components (PCs). The significance threshold for univariate analyses was set at Bonferroni-corrected *P* < 8.33×10^⁻3^ (0.05/6).

### Genetic correlations

Genetic correlational analyses were conducted to explore the shared genetic architecture between CSVD neuroimaging biomarkers and various brain health outcomes using cross-trait LDSC. This method is robust to sample overlap between trait pairs and was implemented using summary statistics from univariate CSVD GWAS, brain health outcome GWAS, and the precomputed LD scores from the European population of the 1000 Genomes Project as the reference panel^62^. Following prior research^63^, the brain health outcomes included a wide range of traits, including clinical diagnoses (MDD^64^, anxiety^65^, ASD^66^, BP^67^, schizophrenia^68^, ADHD^69^, AD^70^, LBD^71^, FTD^72^, ALS^73^, MS^74^, PD^75^ and different types of stroke^76^), general cognitive functions, reaction time and verbal numerical reasoning^77^, fluid intelligence^78^, sleep-related traits (daytime napping^79^, daytime sleepiness^80^, insomnia frequency^81^, sleep duration^79^), risk tolerance^82^, neuroticism^83^, as well as metrics of brain structure (e.g., total brain volume^84^, CSF p-tau levels^85^, and white matter hyperintensities^86^). As recommended, LDSC analyses were restricted to Hapmap3 SNPs, excluding the major histocompatibility complex (MHC) region to avoid spurious associations.

### Bidirectional Mendelian randomization (MR)

To further infer potential causal relationships between our six CSVD biomarkers and brain disorders. we performed bidirectional two-sample MR using previously curated GWAS of 17 major neuropsychiatric disorders, which has no sample overlap with UKB participants. For forward MR, the instrumental variables (IVs) of the exposure (i.e., CSVD neuroimaging traits) were selected using the clumping method in PLINK1.9, with a significance threshold of 5 × 10^⁻8^, a LD *r*² of 0.01 and a window size of 1Mb. Potential outliers detected by Cochran’s Q test and Rucker’s Q′ test were excluded. Four MR methods were applied: inverse variance weighted (IVW), MR-Egger, weighted median, and weighted mode, with IVW as the primary method for reporting results. We also conducted MR-PRESSO and MR-Egger analysis for detecting horizontal and directional pleiotropy to ensure the MR assumptions were not violated. All MR analyses were carried out using the TwoSampleMR package in R. Reverse MR analysis was also conducted to exclude reverse causal associations, with brain disorders as exposure and CSVD neuroimaging traits as outcome, and genetic instruments selection and outlier detection performed using the same procedure as the forward MR.

### Colocalization analysis

To examine whether the CSVD biomarkers and brain disorders with significant MR relationships shared the same causal variants, the colocalization analysis was performed using the coloc R package^87^, using the default priors: *P*^1^ *=* 10^−4^*, P*^2^ *=* 10^−4^, and *P*^12^ *=* 10^−5^. The colocalization analysis tested five different hypotheses and we regarded those with a posterior probability PP.H4 ≥ 0.8 as strong evidence for a shared causal variant affecting both CSVD neuroimaging traits and brain disorders.

### Drug target identification

To identified potential drug targets for neuroimaging informed CSVD, we applied two-sample MR between circulating protein levels and CSVD neuroimaging traits. Specifically, both CSF and plasma protein quantitative trait loci (pQTL) were used. CSF pQTL summary statistics were obtained for 7,028 proteins measured by Somascan 7K platform in 3,107 European participants^76^. Plasma pQTL summary statistics were obtained for 2,923 proteins measured by Olink Explore 3072 in 52,363 UKB European participants^52^. In the drug target analysis, we performed the GWAS of CSVD neuroimaging traits on participants with neuroimaging data but without proteomic data, thereby meeting the non-overlapping sample assumption required for two-sample MR. The instrument variants were identified in the following steps^76^: (1) identify cis-pQTL by selecting variants within 1Mb of the transcription start site of the protein coding gene; (2) select independent variants using clumping method of PLINK2, with a *P* < 5 × 10^⁻8^, a LD *r*² of 0.01 and a window size of 1Mb; (3) exclude variants fall within MHC region. Following these steps, we performed two-sample cis-MR between circulating protein levels and CSVD neuroimaging traits. In the forward MR, five MR methods were applied: inverse variance weighted (IVW), MR-Egger, weighted median, weighted mode and Wald ratio, with IVW as the primary method for reporting results and the last one when only one instrument was valid. Reverse MR was conducted to rule out the reverse causation, by using CSVD neuroimaging traits as the exposure and the circulating protein levels as outcome^54,88^. To rule out the potential violation of MR assumptions, we performed sensitivity analyses using MR-PRESSO for detecting horizontal pleiotropy and MR-Egger analysis for directional pleiotropy.

Using the proteins identified in significant *cis*-MR associations, we identified corresponding drugs with potential for repurposing. If the same protein showing opposite direction of effect across CSF and plasma, we retained the *cis*-MR relationship observed in CSF. Specifically, the corresponding drugs were first systemically retrieved from the DGIdb, ChEMBL, pharmGKB, DrugBank and TTD databases, and subsequently supplemented and validated with the up-to-date publications. Drugs that were either approved or in investigation and demonstrated modes of action in opposite to the *cis*-MR estimate, i.e., anti-CSVD, were manually screened. In other words, the drugs with inhibiting effects were eligible for proteins with positive *cis*-MR relationships and those with promoting effects were selected for proteins with negative *cis*-MR relationships.

## Declarations

### Data availability

The imaging, health outcomes, phenotype, plasma proteomic, metabolic, and genetic data are publicly available at the official website of UK Biobank (http://www.ukbiobank.ac.uk/) and were used following the application no. under application number 202239 and 19542. All software and methods used in our study are publicly available. The code to reproduce the results can be accessed at https://github.com/weikanggong1/CSVDtransformer.git.

## Acknowledgments

We want to thank all the participants and researchers from the UK Biobank. W.C. was supported by grants from the Noncommunicable Chronic Diseases-National Science and Technology Major Project (2025ZD0546300), the National Key R&D Program of China (No. 2023YFC3605400), the National Natural Science Foundation of China (No. 82472055, No. 62433008), the Shanghai Pilot Program for Basic Research - Fudan University 21TQ1400100 (25TQ010), and Shanghai Science and Technology Commission Program (23JS1410100). J.-T.Y. was supported by grants from the Science and Technology Innovation 2030 Major Projects (no. 2022ZD0211600), the National Natural Science Foundation of China (no. 82071201, 81971032 and 92249305), the Shanghai Municipal Science and Technology Major Project (no. 2018SHZDZX01), the Research Start-Up Fund of Huashan Hospital (no. 2022QD002), the Excellence 2025 Talent Cultivation Program at Fudan University (no. 3030277001), Shanghai Talent Development Funding for the Project (no. 2019074), and the Zhangjiang Lab, Tianqiao and Chrissy Chen Institute, and the State Key Laboratory of Neurobiology and Frontiers Center for Brain Science of Ministry of Education, Shanghai Academy of Natural Sciences (SANS), Shanghai Medical College, Fudan University. The funders had no role in study design, data collection and analysis, decision to publish or preparation of the manuscript. Several icons were created with BioRender.com.

## Author information

### Contributions

W.G., M.C., J.Y., and W.C. Conceptualization, Supervision, Project administration, Writing-Reviewing and Editing, Funding acquisition. W.G., F,L., P.R. Software, Formal analysis, Hardware, Validation, Data Analysis, Writing-Original draft preparation, Reviewing and Editing. H.H. Hardware, Methodology, Validation, Data Analysis, Reviewing, W.S. Methodology, Formal analysis, Visualization, Editing. H-Y.H., Q.L., L.H., Q.H., Y.Z., Y.F., D.Z., M.M., R.X., T.W., L.Z., S.H., B.L., L.H. Data Analysis, Validation. X.H., W.Z., Z.L. Methodology, Data Analysis, Hardware.

### Supplementary Material

Supplementary material is available at *Nature Medicine* online.

